# Beyond social prescribing - the use of social return on investment (SROI) analysis in integrated health and social care interventions in England and Wales: a protocol for a systematic review

**DOI:** 10.1101/2022.10.28.22281599

**Authors:** Genevieve Hopkins, Eira Winrow, Ceryl Davies, Diane Seddon

**Author notes:** **Contributors** GH conceptualised this study. GH wrote the first draft. EW, CD and DS agreed the methodology, provided feedback on the first draft of this protocol paper, and agreed the final draft. GH and EW will lead the systematic review. **Data availability statement** This protocol does not report data. **Competing interests** None.

## Abstract

**Introduction:** With increasing costs of healthcare in England and Wales following the COVID-19 pandemic, finding alternatives to traditional medical interventions is more important than ever. Social prescribing provides a way of addressing health and well-being through using non-medical methods that may help relieve costs to the NHS.

Evaluating interventions, such as social prescribing, which have high social (but not easily quantifiable) value, can be problematic. Social return on investment (SROI) is a method of assigning monetary values to both social value as well as traditional assets, so provides a way of evaluating social prescribing initiatives.

**Method:** This protocol outlines the steps that will be taken in a systematic review of the SROI literature surrounding social prescribing-type integrated health and social care interventions based in the community in England and Wales.

Online academic databases such as PubMed Central, ASSIA and Web of Science will be searched, as will grey literature sources such as Google Scholar, the Wales School for Social Prescribing Research (WSSPR) and Social Value UK.

Titles and abstracts from the articles returned by the searches will be reviewed by one researcher. Those selected for full text review will be independently reviewed and compared by two researchers. Where the researchers disagree a third reviewer will help resolve any differences.

Information collected will include identifying stakeholder groups, assessing the quality of SROI analyses, identifying intended and unintended changes of social prescribing interventions, and comparing social prescribing initiatives in terms of SROI costs and benefits.

Quality assessment will be independently conducted on the selected papers by two researchers. The researchers will discuss to obtain consensus. Where there is disagreement, a third researcher will resolve these cases. A pre-existing quality framework will be developed and used to assess the quality of the literature.

**Protocol registration:** **Prospero registration number:** CRD42022318911

https://www.crd.york.ac.uk/prospero/display_record.php?ID=CRD42022318911

## Introduction

### Social prescribing

Social prescribing is a method for improving the well-being of individuals. It often addresses a particular issue, such as physical health, but it can often have ongoing positive consequences [1]; examples include, supporting positive mental health, promoting social inclusion, and reducing loneliness[2].

The delivery of social prescribing varies considerably. In England, it typically involves four parties: primary care, a facilitator, the third sector and the beneficiary. It is instigated by a primary care practitioner, who refers people to a facilitator, known as a social prescribing coordinator [3] or link worker [4]. The link worker uses their local knowledge to work with beneficiaries to co-design and co-prescribe services from within third sector community-based or voluntary organisations to address the beneficiaries’ needs [5].

In Wales, people benefiting from social prescribing services have a greater variety of pathways into social prescribing activities [6]. Although, referral through primary care does take place, it typically happens through self-referral to third party organisations or through Local Authorities [6].

At the far end of the social prescribing spectrum from the classic social prescribing model, are integrated health and social care approaches, such as Asset-Based Community Development (ABCD). These approaches view traditional systems as instigated from the top down and as negative in outlook; government agencies determine what is lacking within a community and then seek to fill those perceived gaps. Some maintain that this is not financially viable [7], while others state that it is also unworkable [8]. This is partly because of the constant drain on resources as new areas of need are identified, but also because the model is external to the community, not generated by the community, which makes it unsustainable. ABCD looks to identify strengths, map assets and potential assets within a community and to utilise them [9]. By forging new links between these assets [10] and promoting self-generation from within the community to build capacity [9], ABCD seeks to develop long-term solutions to health and well-being. The role of external agencies is to provide a facilitating role and help fill gaps that the community assets are unable to [8].

### Social prescribing costs and commitment by English and Welsh administrations

Social prescribing is perceived as a cost-saving alternative to medical-based solutions; it is considered to reduce the use of primary care facilities and prescribed medications [11]. Although, this is not in fact true, it does shift some of the burden of cost from primary care to the third sector [12]. That the third sector then needs to be properly funded to be able to deliver to standard [13] is beyond the scope of this review.

This perceived reduction in costs, plus a commitment to holistic approaches, reducing health inequalities and Universal Personal Care has resulted in a policy shift in England to promote the use of social prescribing. For example, the NHS, in their Long-Term Plan, are committed to referring at least 900,000 people to social prescribing by 2022/23 [14].

In Wales, the Welsh Government have a commitment to maintaining and improving the well-being of the residents of Wales. Both the Social Services and Well-being (Wales) Act 2014, and Well-being of Future Generations (Wales) Act 2015 have had ‘a profound impact on how well-being is being understood, enhanced, and promoted across Wales’ [15]. Social prescribing offers a useful mechanism for delivering the goals of these Acts, specifically around the development of preventative public services and promoting overall community well-being. The Welsh Government have explicitly outlined their commitment to social prescribing; they specify the introduction of a Welsh framework for developing social prescribing programmes to help end social isolation [16].

With increasing financial pressures on the UK economy, particularly due to the healthcare costs resulting from the COVID-19 pandemic, alongside the commitment to social prescribing from the English and Welsh administrations, it is important that reliable methods of evaluation are used to measure the cost-benefits of social prescribing.

### The costs of healthcare and COVID-19

Nominal healthcare spending was relatively consistent in the UK in the ten years between 2009 and 2019 and accounted for around 10 per cent of GDP [17]. However, expenditure in the UK rose by 20 per cent between 2020 and 2021 to around 12 per cent of GDP [17]. This was the largest year-on-year rise in healthcare spending since comparative records began in 1997. The rise was due to the global COVID-19 pandemic, which had two contributing effects. Firstly, although some aspects of healthcare spending decreased (for example, a reduction in scheduled primary care and hospital appointments [18]), overall spending increased due to factors such as the rise in use of personal protective equipment (£14.8 billion) and the introduction of the Test and Trace programme (£23 billion) between 2020 and 2021 [19]. Secondly, GDP itself decreased because of the drop in UK economic output [20]. For the year 2021/22, the NHS budget was £192 billion, with an estimated £33.8 billion of this used to tackle the COVID-19 pandemic [21].

Looking at spending in primary healthcare, more than 20 per cent of people attend a GP’s surgery for non-medical reasons [22]; the average GP appointment lasts 9 minutes and costs £39.23 [23]. This puts a substantial time and cost burden on the UK primary care system and leads GPs to be most likely to recommend and overprescribe drug-based interventions, such as antidepressants [24,25]. Prior to the Covid-19 pandemic, depression was present in around 10 per cent of the adult population of Great Britain [26]; this increased to 21 per cent in between January and March 2021 then decreased to 17 per cent by summer 2021. This was reflected in spending on antidepressants which in the year before the pandemic (2019/2020) cost £226 million in England [27] and £16 million in Wales [28], rose to £375 million in England [27] and £28 million in Wales [28], during the pandemic (2020/2021) and dropped to £247 million in England [27] and £18 million in Wales [28], following the main period of the pandemic (2021/2022).

### SROI

Social Return on Investment (SROI) is a principles-based method of cost-benefit analysis (CBA) used to assign monetary values to assets or outcomes (economic, environmental, or social) that are not generally accounted for in standard financial accounts to determine a project or intervention’s social value [29].

The aim is to include the values of people that are often excluded from markets in the same terms as used in markets, to give people a voice in resource allocation decisions along with narrative types of information about value [30]. SROI allows researchers to identify what changes and what is important for individuals, organisations and any other stakeholders, giving a much wider measure of value.

### What this systematic review will do

The paper outlines a protocol of the methodology that will be used to review the relevant literature on the use of SROI analysis for social prescribing initiatives.

### Social return on investment methodology

SROI is governed and promoted through Social Value UK and Social Value International. These organisations are a network of certified professionals that promote SROI as a method to account for social value and the way it is evaluated. SROI methodology, which originated in the US in the mid-90s and was subsequently adopted in the UK following a government-funded initiative to measure social value in 2008 [31] continues to evolve [32].

To calculate the social value generated by an intervention, an impact map is created during SROI analysis. This involves six stages [33]:

1. Identifying key stakeholders – anyone affected by the programme
2. Mapping outcomes – what changes and for whom – this encompasses theory of change
3. Giving outcomes a value – measuring outcomes and applying proxy values where necessary. Social Value UK and Housing Associations’ Charitable Trust (HACT) have produced a Social Value Bank that can be used to assign proxy values for outcomes. It contains 53 outcomes that are sub-divided by age and geography to produce 636 values. Areas that the Bank covers include employment, local environment, health, financial inclusion, housing, social groups and physical activity [34].
4. Establishing impact – accounting for attribution, deadweight, and drop-off – these are adjustments made to ensure that social value is not ‘over-claimed’; this is one of the seven principles of SROI – [35]. Attribution is the amount of change due to other activities external to the intervention, deadweight is amount of change that would have happened if the intervention had not taken place, while drop-off is the amount of change that drops of after the first year.
5. Calculating the SROI -the SROI ratio is calculated by dividing the total value of outcomes by the total value of inputs across all stakeholders. The SROI ratio is the amount of social value generated for every £1 invested in the programme or intervention. The base or best-case scenario is usually reported, following a range of sensitivity analyses to explore how the SROI ratio would be affected if parameters were changed, different financial proxies were used, and different levels of outcomes were observed to those demonstrated in the base case or final reported SROI ratio.
6. Reporting, using, and embedding – sharing of results of the SROI analysis with stakeholders and responding to their feedback.

There are two types of SROI analysis: evaluative, which is conducted retrospectively and based on outcomes that have already taken place and forecast, which predicts how much social value will be created if the activities meet their intended outcomes [36]. This systematic review will examine both evaluative and forecast SROI evidence.

### Previous systematic reviews of SROI and social prescribing

Six previous, related SROI reviews have taken place [33,36-40]. One conducted a meta-analysis of SROI on ‘short’ documents (6 to 90 pages) [37]. Search terms included SROI and similar terms such as social impact analysis and social benefit analysis; there were no restrictions on the type of intervention or the organisations providing the interventions.

Of the remaining five reviews, four looked at public health interventions [33, 38-40] while only one looked specifically at social prescribing-type interventions and focussed on physical and sport activities [36].

The reviews concentrated on high-income countries, or their search criteria returned a high number of studies from high income countries, and except for one that looked both at peer-reviewed and grey literature [39].

This will be the first systematic review to look solely at SROI analysis of integrated health and social care social prescribing activities delivered in community settings, in the England and Wales that have been reported both in peer-reviewed and grey literature.

### The current study

The purpose of this systematic review is to summarise studies that use SROI to evaluate social prescribing type schemes to:

1. identify stakeholder groups that typically contribute to SROI analyses in social prescribing
2. assess the quality of SROI analysis in social prescribing studies
3. identify intended/unintended changes of social prescribing interventions in SROI analyses
4. compare social prescribing initiatives in terms of SROI costs and benefits

## Methods

### Design

The review will be conducted using the Preferred Reporting Items for Systematic Reviews and Meta-Analyses (PRISMA) checklist and flow diagram [41] for evaluating interventions as guidance. This protocol is registered with the International Prospective Register of Systematic Reviews, PROSPERO (number CRD42022318911)

### Inclusion criteria

Studies which contain an SROI evaluation of a social prescribing-type interventions. The focus is on integrated health and social care interventions delivered in the community; that is programmes that are designed to address both elements of health (either mental and physical) and well-being, that are non-drug interventions based in non-institutional settings in the England and Wales. Peer-reviewed publications and those classed as ‘grey’ literature will be considered for inclusion.

### Exclusion criteria

Only studies involving adults are eligible, as this review will inform an evaluation of an integrated health and social care social prescribing model for adults aged 18 years and over. Other forms of financial analysis of interventions in social prescribing apart from SROI are not considered. Publications that are not written in English will not be considered.

### Types of studies

All studies that have an SROI component for social prescribing initiatives for adult groups or adult populations.

### Search strategy

Documentation in English and Welsh over the last 20 years (2002 to 2022) will be screened. Searching will take place using the online databases PubMed Central, ASSIA, Web of Science, CINAHL, the Cochrane Library, Medline via Ovid, PyscInfo. As much SROI literature is grey literature [37, 42], other sources will be searched, including: Google, Google Scholar, NHS England, Wales School for Social Prescribing Research (WSSPR), Social Prescribing Network, Social Value UK and Social Value International.

Search terms are to be specific and focussed as this is intrinsic to the review; they have been informed by other systematic reviews of social prescribing. As SROI is being targeted, no synonyms will be used to substitute for it. The search terms to be used are included in the supporting file (S1).

Extraction of documentation will be conducted using PICO terms (Population, Intervention, Comparator, Outcomes); the participants will be adults, the intervention, social prescribing interventions that take place in the community that have been assessed using SROI analysis, with any or no comparator and the main outcome(s) improved health and well-being

### Screening and data extraction

Following extraction using PICO terms into Mendeley Reference Manager, the titles of extracted articles will be matched against inclusion and exclusion criteria. Those that meet the criteria will have their abstracts matched against the criteria. Articles with abstracts matching the criteria will have their full articles retrieved. Articles that do not meet the inclusion criteria will be hand sifted for other relevant articles, and the title, abstracts and full texts of the other relevant articles will be compared against the inclusion and exclusion criteria. The number of articles that do not meet the inclusion criteria will be recorded in a PRISMA diagram [41], as will the reasons for their exclusion.

The titles and abstracts will be reviewed by GH, and those selected for full text review will be independently reviewed and compared by two different researchers (GH, EW). Where the researchers disagree whether to include or exclude articles, (CD or DS) will help to resolve these cases in discussion with researchers GH and EW.

The data extracted from the selected articles will be recorded in an Excel table; the rows will represent the articles, and the columns will represent the data fields to be extracted. The data fields will form two main categories: those that provide descriptive data on the articles and those that speak to the validity of the data (see section on risk of bias). Categories providing descriptive data will include author(s), title of article, date article published, journal/publication site, country, type of participants (age, sex, gender, ethnicity, rurality, socio-economic status, occupation, health status), interventions, comparators, outcome measures and types of study.

Categories providing SROI data will include stakeholder groups, intended/unintended changes of social prescribing interventions, value of inputs (costs), value of benefits, base SROI ratio, sensitivity analysis estimate lower ratio and sensitivity analysis estimate upper ratio.

### Quality assessment

Quality assessment will be independently conducted on the selected papers by two researchers (GH and EW). The researchers will discuss to obtain consensus. Where there is disagreement, a third researcher (CD or DS) will resolve these cases.

A 21-point SROI Quality Framework [42] will be used as a basis to assess the quality of the documents meeting the inclusion criteria. The Framework was developed from, and supplements criteria developed in previous studies [33, 37]. It has the benefit of containing a comprehensive range of SROI-specific measures, including justification of the method, the level of contribution of stakeholders, participant consent, clarity of theory of change and the contribution of intended and unintended outcomes, appropriate study design and sampling, the inclusion of valid and comprehensive financial and non-financial inputs, and accounting for and justifying deadweight, sensitivity analysis and limitations. Scoring for each element is of the mutually exclusive categories ‘yes’, ‘no’, ‘not clear’ and ‘not applicable’. Articles score one point for each ‘yes’ on the Framework.

Some of the elements of the Framework have dual requirements [39], for example, one element is ‘Was dead-weight clearly described and calculated?’; requires both the description and calculation of dead-weight to be present to fulfil the criterion, but only gains one point in the scoring system. Consequently, the framework will be expanded to account for this.

### Data synthesis

A narrative synthesis will be performed to establish the SROI features of each paper, and tables will be used to identify stakeholder groups and intended/unintended changes of social prescribing interventions in SROI analyses. An assessment of the quality of SROI analysis in the studies will be undertaken as described above.

### Analysis of subgroups or subsets

None

### Patient and public involvement

As this protocol outlines the process of a systematic review, no direct patient or public involvement was sought.

## Supporting information

S1 Supplemental file - search strategy

PRISMA checklist

## Data Availability

No datasets were generated or analysed during the current study. All relevant data from this study will be made available upon study completion.

## Ethics and dissemination

This paper describes the protocol for a systematic review of literature in SROI in social prescribing. As a protocol, it does not require ethical approval. The results of the systematic review will be disseminated through a peer-reviewed journal article.

## Supporting information

S1 Systematic review search strategy

## References

1. Ellis-Hill C, Thomas S, Gracey F, Lamont-Robinson C, Cant R, Marques E et al. HeART of Stroke: randomised controlled, parallel-arm, feasibility study of a community-based arts and health intervention plus usual care compared with usual care to increase psychological well-being in people following a stroke. BMJ Open. 2019;9(3):e021098. doi: 10.1136/bmjopen-2017-021098

2. Wakefield J, Bowe M, Kellezi B, Butcher A, Groeger J. Longitudinal associations between family identification, loneliness, depression, and sleep quality. British Journal of Health Psychology. 2019;25(1):1–16. doi: 10.1111/bjhp.12391

3. Bertotti M, Frostick C, Hutt P, Sohanpal R, Carnes D. A realist evaluation of social prescribing: an exploration into the context and mechanisms underpinning a pathway linking primary care with the voluntary sector. Primary Health Care Research & Development. 2017;19(03):232–245. doi: 10.1017/s1463423617000706

4. Wildman J, Moffatt S, Steer M, Laing K, Penn L, O’Brien N. Service-users’ perspectives of link worker social prescribing: a qualitative follow-up study. BMC Public Health. 2019;19(1). doi: 10.1186/s12889-018-6349-x

5. Husk K, Blockley K, Lovell R, Bethel A, Lang I, Byng R et al. What approaches to social prescribing work, for whom, and in what circumstances? A realist review. Health & Social Care in the Community. 2019;28(2):309–324. doi: 10.1111/hsc.12839

6. Rees S, Carolyn W, Davies M. Creating sustainable community assets/social capital within the context of social prescribing: Findings from the workshop held 17/07/19. Wsspr, wales. 2019. [accessed 7 Oct 2022] Available from: http://www.wsspr.wales/resources/Rees%20et%20al%202019.pdf

7. Lamb J, Dowrick C, Burroughs H, Beatty S, Edwards S, Bristow K et al. Community Engagement in a complex intervention to improve access to primary mental health care for hard-to-reach groups. Health Expectations. 2014;18(6):2865–2879. doi: 10.1111/hex.12272

8. Russell C. Communities-in-Control-Developing-Assets.pdf. Academia.edu. 2009. [accessed 7 Oct 2022] Available from: https://www.academia.edu/37616739/Communities_in_Control_Developing_Assets_pdf

9. Hufford L, West D, Paterniti D, Pan R. Community-Based Advocacy Training: Applying Asset-Based Community Development in Resident Education. Academic Medicine. 2009;84(6):765–770. doi: 10.1097/acm.0b013e3181a426c8

10. Kretzman J, McKnight J. Building communities from the inside out. Vancouver, B.C.: Langara College; 2004.

11. Dixon M, Polley M. Report of the annual social prescribing network conference. 2016. [accessed 7 Oct 2022] Available from: https://www.researchgate.net/publication/359393191_REPORT_OF_THE_ANNUAL_SOCIAL_PRESCR IBING_NETWORK_CONFERENCE

12. South J, Higgins TJ, Woodall J, White SM. Can social prescribing provide the missing link? Primary Health Care Research & Development. Cambridge University Press; 2008;9(4):310–8.

13. Islam M. Social Prescribing—An Effort to Apply a Common Knowledge: Impelling Forces and Challenges. Frontiers in Public Health. 2020;8. doi: 10.3389/fpubh.2020.515469

14. NHS. The NHS Long Term Plan. NHS Long Term Plan. 2019. [accessed 7 Oct 2022] Available from: https://www.longtermplan.nhs.uk/publication/nhs-long-term-plan/

15. Wallace C, Davies M, Elliott M, Llewellyn M, Randall H, Owens J et al. Understanding Social Prescribing in Wales: A Mixed Methods Study. University of South Wales. 2022. [accessed 7 Oct 2022] Available from: https://pure.southwales.ac.uk/en/publications/understanding-social-prescribing-in-wales-a-mixed-methods-study

16. Welsh Government. Developing a national framework for social prescribing | GOV.WALES. GOV.WALES. 2022. [accessed 7 Oct 2022] Available from: https://gov.wales/developing-national-framework-social-prescribing

17. ONS. Healthcare expenditure, UK Health Accounts provisional estimates - Office for National Statistics. Ons.gov.uk. 2022. [accessed 7 Oct 2022] Available from: https://www.ons.gov.uk/peoplepopulationandcommunity/healthandsocialcare/healthcaresystem/b ulletins/healthcareexpenditureukhealthaccountsprovisionalestimates/2020

18. Propper C, Stoye G, Zaranko B. The Wider Impacts of the Coronavirus Pandemic on the NHS *. Fiscal Studies. 2020;41(2):345–356. doi: 10.1111/1475-5890.12227

19. DHSC. DHSC annual report and accounts: 2020 to 2021. GOV.UK. 2022. [accessed 7 Oct 2022] Available from: https://www.gov.uk/government/publications/dhsc-annual-report-and-accounts-2020-to-2021

20. ONS. Healthcare expenditure, UK Health Accounts provisional estimates - Office for National Statistics. Ons.gov.uk. 2021. [accessed 7 Oct 2022] Available from: https://www.ons.gov.uk/peoplepopulationandcommunity/healthandsocialcare/healthcaresystem/b ulletins/healthcareexpenditureukhealthaccountsprovisionalestimates/2020

21. King’s Fund. Key facts and figures about the NHS. 2022. [accessed 7 Oct 2022] Available from: https://www.kingsfund.org.uk/audio-video/key-facts-figures-nhs

22. Torjesen I. Social prescribing could help alleviate pressure on GPs. BMJ. 2016; i1436. doi: 10.1136/bmj.i1436

23. Curtis L, Burns A. Unit Costs of Health and Social Care 2020 | PSSRU. Pssru.ac.uk. 2020. [accessed 7 Oct 2022] Available from: https://www.pssru.ac.uk/project-pages/unit-costs/unit-costs-2020/

24. Spence D. Are antidepressants overprescribed? Yes. BMJ. 2013;346(jan22 3):f191–f191. doi: 10.1136/bmj.f191

25. McCartney M. Overprescribing antidepressants: where’s the evidence?. BMJ. 2014;348(jun30 1):g4218-g4218. doi: 10.1136/bmj.g4218

26. UK Government. Coronavirus and depression in adults, Great Britain: July to August 2021. GOV.UK. 2021. [accessed 7 Oct 2022] Available from: https://www.gov.uk/government/statistics/coronavirus-and-depression-in-adults-great-britain-july-to-august-2021

27. NHSBSA Medicines Used in Mental Health – England – Quarterly Summary Statistics January to March 2022 | NHSBSA. Nhsbsa.nhs.uk. 2022. [accessed 7 Oct 2022] Available from: https://www.nhsbsa.nhs.uk/statistical-collections/medicines-used-mental-health-england/medicines-used-mental-health-england-quarterly-summary-statistics-january-march-2022

28. Welsh Government. Prescriptions in Wales: April 2021 to March 2022 | GOV.WALES. GOV.WALES. 2022. [accessed 7 Oct 2022] Available from: https://gov.wales/prescriptions-wales-april-2021-march-2022

29. Willis E, Semple A, de Waal H. Quantifying the benefits of peer support for people with dementia: A Social Return on Investment (SROI) study. Dementia. 2016;17(3):266–278. doi: 10.1177/1471301216640184

30. Charles JM, Jones A, Lloyd-Williams H. In: Edwards R, McIntosh E. editors. Applied health economics for public health practice and research. Oxford University Press; 2019. pp 279–300.

31. Arvidson M, Lyon F, McKay S, Moro D. Valuing the social? The nature and controversies of measuring social return on investment (SROI). Voluntary Sector Review. 2013;4(1):3–18. doi: 10.1332/204080513x661554

32. Social Value UK. About Us - Social Value UK. Social Value UK. 2022. [accessed 7 Oct 2022] Available from: https://socialvalueuk.org/about-social-value-uk/

33. Banke-Thomas A, Madaj B, Charles A, van den Broek N. Social Return on Investment (SROI) methodology to account for value for money of public health interventions: a systematic review. BMC Public Health. 2015;15(1). doi: 10.1186/s12889-015-1935-7

34. Social Value UK/HACT. SROI and HACT Social Value Bank Linkages Paper Social Value UK. Social Value UK 2015. [accessed 7 Oct 2022] Available from: https://socialvalueuk.org/resource/sroi-and-hact-social-value-bank-linkages-paper/

35. Fujiwara D. The Seven Principle Problems of SROI. 2015. [accessed 7 Oct 2022] Available from: https://www.semanticscholar.org/paper/The-Seven-Principle-Problems-of-SROI-Fujiwara/a3ff4a7770a8f71a82d55ed42aefcd2138de0faf

36. Gosselin V, Boccanfuso D, Laberge S. Social return on investment (SROI) method to evaluate physical activity and sport interventions: a systematic review. International Journal of Behavioral Nutrition and Physical Activity. 2020;17(1). doi: 10.1186/s12966-020-00931-w

37. Krlev G, Munscher R, Mulbert K. Social Return on Investment (SROI): state-of-the-art and perspectives a meta-analysis of practice in Social Return on Investment (SROI) studies published 2002-2012. 2013. [accessed 7 Oct 2022] Available from: https://www.researchgate.net/publication/280664441_Social_Return_on_Investment_SROI_state-of-the-art_and_perspectives_-_a_meta-analysis_of_practice_in_Social_Return_on_Investment_SROI_studies_published_2002-2012

38. Masters R, Anwar E, Collins B, Cookson R, Capewell S. Return on investment of public health interventions: a systematic review. Journal of Epidemiology and Community Health. 2017;71(8):827–834. doi: 10.1136/jech-2016-208141

39. Hutchinson C, Berndt A, Forsythe D, Gilbert-Hunt S, George S, Ratcliffe J. Valuing the impact of health and social care programs using social return on investment analysis: how have academics advanced the methodology? A systematic review. BMJ Open. 2019;9(8):e029789. doi: 10.1136/bmjopen-2019-029789

40. Ashton K, Schröder-Bäck P, Clemens T, Dyakova M, Stielke A, Bellis M. The social value of investing in public health across the life course: a systematic scoping review. BMC Public Health. 2020;20(1). doi: 10.1186/s12889-020-08685-7

41. Page M, McKenzie J, Bossuyt I, Boutron I, Hoffmann T, Mulrow C et al. The PRISMA 2020 statement: an updated guideline for reporting systematic reviews. 2021. [accessed 7 Oct 2022] Available from: https://pubmed.ncbi.nlm.nih.gov/33782057/

42. Hutchinson C, Berndt A, Gilbert-Hunt S, George S, Ratcliffe J. Valuing the impact of health and social care programmes using social return on investment analysis: how have academics advanced the methodology? A protocol for a systematic review of peer-reviewed literature. BMJ Open. 2018;8(12):e022534. doi: 10.1136/bmjopen-2018-022534

