## Supplementary material for "Beyond social prescribing - the use of social return on investment (SROI) analysis in integrated health and social care interventions in England and Wales: a protocol for a systematic review": S1 Supplemental file - search strategy

The search terms that will be used are:

|  | social prescribing |
| --- | --- |
| OR | social referral |
| OR | social intervention |
| OR | non-medical intervention |
| OR | community prescribing |
| OR | community referral |
| OR | non-medical referral |
| OR | signposting referral |
| OR | information referral |
| OR | wellbeing program |
| OR | well-being program |
| OR | community-based intervention |
| OR | health and wellbeing intervention |
| OR | health and well-being intervention |
| AND | social return on investment |
| OR | SROI |

The synonyms for social prescribing have been informed by the following documents/studies:

Hutchinson C, Berndt A, Gilbert-Hunt S, George S, Ratcliffe J. Valuing the impact of health and social care programmes using social return on investment analysis: how have academics advanced the methodology? A protocol for a systematic review of peer-reviewed literature. BMJ Open. 2018;8(12):e022534. doi: 10.1136/bmjopen-2018-022534

Kiely B, Clyne B, Boland F, et al. Link workers providing social prescribing and health and social care coordination for people with multimorbidity in socially deprived areas (the LinkMM trial): protocol for a pragmatic randomised controlled trial. BMJ Open 2021;11:e041809. doi:10.1136/ bmjopen-2020-041809

Reinhardt G, Vidovic D, Hammerton C. Understanding loneliness: a systematic review of the impact of social prescribing initiatives on loneliness. Perspectives in Public Health. 2021;141(4):204-213. doi:10.1177/1757913920967040

Bickerdike L, Booth A, Wilson PM*, et al*

Social prescribing: less rhetoric and more reality. A systematic review of the evidence

*BMJ Open* 2017;**7:**e013384. doi: 10.1136/bmjopen-2016-013384

Thomas G, Lynch M, Spencer LH. A Systematic Review to Examine the Evidence in Developing Social Prescribing Interventions That Apply a Co-Productive, Co-Designed Approach to Improve Well-Being Outcomes in a Community Setting. Int J Environ Res Public Health. 2021 Apr 8;18(8):3896. doi: 10.3390/ijerph18083896. PMID: 33917681; PMCID: PMC8067989.

Oluseyi Florence Jimoh, Chris Fox, Toby Smith, Jane Fox, Euan Sadler, Mizanur Khondoker, Jenny Whitty, Alan Louise, Anne Corbett, Jose Valderas Martinez. Effectiveness of social prescribing interventions to prevent or delay frailty in community living older adults: a systematic review of the evidence. PROSPERO 2019 CRD42019141868

Mark Davies, Carolyn Wallace, David Pontin, Sarah Wallace. Enhancing student wellbeing through social prescribing - a rapid realist review. PROSPERO 2020 CRD42020193075
